# Geographic reconstruction of the SARS-CoV-2 outbreak in Lombardy (Italy) during the early phase

**DOI:** 10.1101/2020.07.23.20159871

**Authors:** Micheli Valeria, Rimoldi Sara Giordana, Romeri Francesca, Comandatore Francesco, Mancon Alessandro, Gigantiello Anna, Brilli Matteo, Mileto Davide, Pagani Cristina, Lombardi Alessandra, Gismondo Maria Rita, Laboratory of Clinical Microbiology, Virology and Diagnostic of Bioemergencies group

**Affiliations:** Laboratory of Clinical Microbiology, Virology and Diagnostic of Bioemergencies, ASST Fatebenefratelli Sacco, Milan, Italy. G.B. Grassi 74, 20157 Milan, Italy; Romeo ed Enrica Invernizzi Pediatric Research Center, Department of Biomedical and Clinical Sciences L. Sacco, University of Milan, Italy

**Keywords:** SARSCoV-2, Lombardy Region, WGS

## Abstract

The circulation of SARS-CoV-2 in Italy has been dominated by two large clusters of outbreaks in Northern part of the peninsula, source of alarming and prolonged infections in Lombardy region, in Codogno and Bergamo areas especially.

The aim of the study was to expand understanding on the circulation of SARS-CoV-2 in the affected Lombardy areas. To this purpose, twenty full length genomes were collected from patients addressing to several Lombard hospitals from February 20^th^ to April 4^th^, 2020.

The obtained genome assemblies, available on the GISAD database and performed at the Referral Center for COVID-19 diagnosis, identified 2 main monophyletic clades, containing 9 and 52 isolates, respectively.

The molecular clock analysis estimated a clusters divergence approximately one month before the first patient identification, supporting the hypothesis that different SARS-CoV-2 strains spread all over the world at different time, but their presence became evident only in late February along with Italian epidemic emergence.

Therefore, the epidemiological reconstruction carried out by this work highlights multiple inputs of the virus into its initial circulation in Lombardy Region.

However, a phylogenetic reconstruction robustness will be improved when other genomic sequences will be available, in order to guarantee a complete epidemiological surveillance.

## INTRODUCTION

In late December 2019, the Chinese health authorities informed the local World Health Organization (WHO) office about the recognition of a pneumonia outbreak of unknown aetiology in Wuhan municipality.^1^ Thanks to the use of Next Generation Sequencing approach, a new coronavirus was rapidly identified as the causative agent of the new Coronavirus Disease 2019 (COVID-19);^2,3^ moreover, due to the rapid global diffusion, the WHO declared the pandemic status on March 11, 2020.^4^

The positive-strand RNA virus family Coronaviridae includes 2 subfamilies for a total of 5 genera and more than 40 species, which infect a broad spectrum of vertebrates;^5^ in particular, seven coronaviruses are related to human diseases, among which two represented a public health concern in the past 20 years: in November 2002, the Severe Acute Respiratory Syndrome related coronavirus (SARS-CoV) emerged in Guangdong, China, resulting in more than 8,000 confirmed cases and 774 deaths in 37 countries; similarly, starting from 2012 in Saudi Arabia, the Middle East Respiratory Syndrome related coronavirus (MERS-CoV) infected 2,494 individuals and caused 858 fatalities.^6, 7^ The COVID-19 related virus was classified as a β-betacoronavirus and, considering its close correlation to SARS-CoV, it was renamed SARS-CoV-2.^8^

In Europe, Italy is one of the most affected areas, accounting for more than 230,000 cases on June 5^th^, 2020.^9^ Northern Italy has reported the highest prevalence in the country, especially in Milano, Brescia and Bergamo provinces, which registered more than 23,000, 14,000 and 13,000 cases^9^, respectively and an infection rate (case divided by resident population) almost double (0.58 and 0.52%, respectively) compared with the rest of Italy (0.32%). In particular, Bergamo and Brescia provinces had to face a high percentage of severe clinical cases presenting an enormous rate of mortality.

The first identification of autochthonous transmission of SARS-CoV-2 in Italy was documented by the Laboratory of Clinical Microbiology, Virology and Bioemergency of L. Sacco University Hospital (Milano, Italy) on 20^th^ February 2020: the patient was a 38 years old male, who was found positive for pneumonia at the Codogno Hospital, without any evidence of epidemiological linkage with COVID-19 cases; soon after the laboratory received thousands of respiratory samples for the confirmation of suspected COVID-19 patients from many regional institutions (Bergamo, Brescia, Cremona, Codogno, Lodi, Milano): owing to the geographical distribution of these specimens, viral sequence data could give insight into SARS-CoV-2 molecular epidemiology and possible local virus introduction.

The aim of the present study was to assess the potential presence of different viral clusters belonging to the six main provinces involved in Lombardy COVID-19 cases in order to highlight peculiar province-dependent viral characteristics.

## MATERIALS AND METHODS

### Study population

The study included SARS-CoV-2 positive samples collected at the Laboratory of Clinical Microbiology, Virology and Bioemergency of L. Sacco University Hospital, referral centre for COVID-19 diagnosis; all processed specimens were either nasopharyngeal swabs or bronchoalveolar lavage fluid (BALF). To try to optimize sequencing performance, only samples positive for all RT-PCR targets (E gene, N gene, ORF1ab; Novel Coronavirus (2019-nCoV) Real Time Multiplex RT-PCR Kit, Liferiver™) and with a Ct value ≤25 were considered. Since the laboratory received samples from other hospitals widely distributed on the regional territory, patients location was highly variable: selection was therefore performed aiming at maximize geographical representation and detection of related diversity. All relevant clinical, demographic and geographical data were recorded.

### RNA viral extraction and reverse-transcription

Total RNA was extracted from 200 μL of primary sample and eluted in 100 μL by means of the QIAMP VIRAL RNA mini kit (Qiagen, Hilden, Germany), according to manufacturer’s instructions. In order to optimize pre-sequencing PCR, the quality, quantity and purity of the genomic RNA were determined using Qubit 4 fluorometer (Thermofisher Scientific Inc, Italy), according to manufacturer’s instructions. cDNA was then synthesized using ImProm-II™ Reverse Transcriptase (RT) and related reagents (Promega Corporation, Italia). The reaction mixture was prepared adding, for each sample: 6 μL of ImProm-II™ 5X Reaction Buffer, 1.8 μL of 5 mM Mg2+, 1 μL of 10 mM Random Primers, 1 μL of 5mM dNTPs, 1 μL of RT, 0.5 μL of RNasin (40 U/μL) and 20 μL of purified RNA, for a total volume of 31.3 μL; reverse-transcription was performed at the following conditions: 37° C for 45 min, 80° C for 5 min.

### Whole genome sequencing

cDNA was amplified using two SARS-CoV-2 specific primers sets in different reaction tubes, in order to obtain multiple fragments covering the whole viral genome. After pooling the two reaction mixtures, DNA was digested, ligated to barcodes, purified and amplified again; a second purification was performed before dsDNA quantification on Qubit 4 and library preparation on One Touch™ 2 instrument (Thermofisher Scientific, Monza, Italy); the One Touch ES instrument (Thermofisher Scientific, Monza, Italy) was used for final enrichment and Ion Torrent™ Personal Genome Machine™ (PGM) System (Thermofisher Scientific, Monza, Italy) for sequencing. All procedures were executed following manufacturer’s instructions.

### Genome assembly

For each sample, the genome assembly was obtained using a mapping-based approach, as follows. Low quality reads bases were trimmed out using Trimmomatic software,^10^ using thirteen different parameter sets (LEADING:3 TRAILING:3 SLIDINGWINDOW:4:15 MINLEN:36, LEADING:3 TRAILING:3 SLIDINGWINDOW:4:20 MINLEN:36, LEADING:3 TRAILING:3 SLIDINGWINDOW:4:25 MINLEN:36, LEADING:3 TRAILING:10 SLIDINGWINDOW:4:15 MINLEN:36, LEADING:3 TRAILING:10 SLIDINGWINDOW:4:20 MINLEN:36, LEADING:3 TRAILING:10 SLIDINGWINDOW:4:25 MINLEN:36, LEADING:3 TRAILING:20 SLIDINGWINDOW:4:15 MINLEN:36, LEADING:3 TRAILING:20 SLIDINGWINDOW:4:20 MINLEN:36, LEADING:3 TRAILING:20 SLIDINGWINDOW:4:25 MINLEN:36, MAXINFO:50:0.3, MAXINFO:50:0.5, MAXINFO:50:0.7, MAXINFO:50:0.9). Then, SNP calling was performed following the GATK Best Practice procedure^11^, using the Wuhan-Hu-1 strain genome (accession MN908947.3) as reference. The genome consensus sequence was obtained on the basis of the identified SNPs and the reference sequence. Reference bases were called in conserved positions with coverage above five, otherwise N were introduced. The procedure produced thirteen genome assemblies per sample, corresponding to the thirteen different trimming parameter sets. For each sample, the genome assembly with the lowest number of undetermined bases (N) as the final genome assembly was selected.

### Molecular clock analysis

A dataset of 41 genome assemblies of Sars-Cov-2 strains isolated in Italy between 20 February 2020 and 30 March 2020 were retrieved from GISAID database^12^, (see Supplementary Table 1 for details). A global dataset including these 41 GISAID (Table 1) genome assemblies and the 20 genome assemblies produced in this study was produced and aligned using MAFFT.^13^

**Table X.** Geographical origin of samples collected and processed at Sacco Hospital. The table summarises the locations of possible transmission for patients enrolled at Sacco Hospital, expressed as provinces, since some patient was already hospitalized, suggesting a nosocomial acquisition.

| Geographical origin | N |
| --- | --- |
| Milano | 6 |
| Lodi | 4 |
| Brescia | 2 |
| Bergamo | 3 |
| Other | 5 |

The low quality alignment regions at the extremities of the alignment were removed using Gblocks with default parameters.^14^ The obtained alignment was subjected to Maximum Likelihood phylogenetic analysis using RaxML.^15^ The obtained tree was then analysed using TempEst^16^ to assess temporal signal. The Hasegawa-Kishino-Yano model (HKY) was found as the simplest evolutionary model using JmodelTest 2.1.10.^17^ Phylogenetic analysis was performed using a Bayesian Markov Chain Monte Carlo (MCMC) method implemented in BEAST, v.1.10.4^18^ with 10 million states and sampling every 1,000 steps. The coalescent priors constant population size and exponential growth were tested for relaxed molecular clock using Path Sampling (PS) and Stepping Stone (SS) sampling.^19^ Then the phylogenetic analysis was repeated with the selected coalescent priori with 100 million states and sampling every 1,000 steps. The convergence of MCMC chain was checked using Tracer v.1.7.1.^20^ The maximum clade credibility (MCC) trees were obtained from the tree posterior distribution using TreeAnnotator (http://beast.community/index.html).

### Ethical approval

All data used in the study were previously made anonymous, according to the requirements set by Italian Data Protection Code (Legislative Decree 196/2003) and the general authorizations issued by the Data Protection Authority. Under Italian law, all sensitive data were deleted and only age, gender and sampling date were collected providing Ethics Committee approval unnecessary (Art. 6 and Art. 9 of Legislative Decree 211/2003).

## RESULTS

### Study population

A total of 20 samples from different subjects were successfully sequenced and included in phylogenetic analysis, attributing the progressive ID HSacco-N (from HSacco-2 to HSacco-21). All patients were resident in Lombardy Region, distributed in different provinces, as reported in table X: in particular, the category ‘Milano’ contains also patients hospitalized for non-COVID-19 disease, for whom SARS-CoV-2 hospital acquisition was supposed; in addition, HSacco-20, in ‘Other’ category, was a nurse living in Bergamo and working at Lodi Hospital.

### Phylogenetic and molecular clock analysis

The obtained 20 Sars-Cov-2 genome assemblies are available on the GISAD database (see Table 2 for details). The comparison of the marginal likelihoods of constant and exponential coalescent models under a log-normal relaxed clock showed that the best fitting model was the exponential coalescent prior (PS BF exponential growth vs. constant = 2,42; SS BF exponential growth vs. constant = -2,39). The obtained phylogenetic tree is reported in Figure 1.

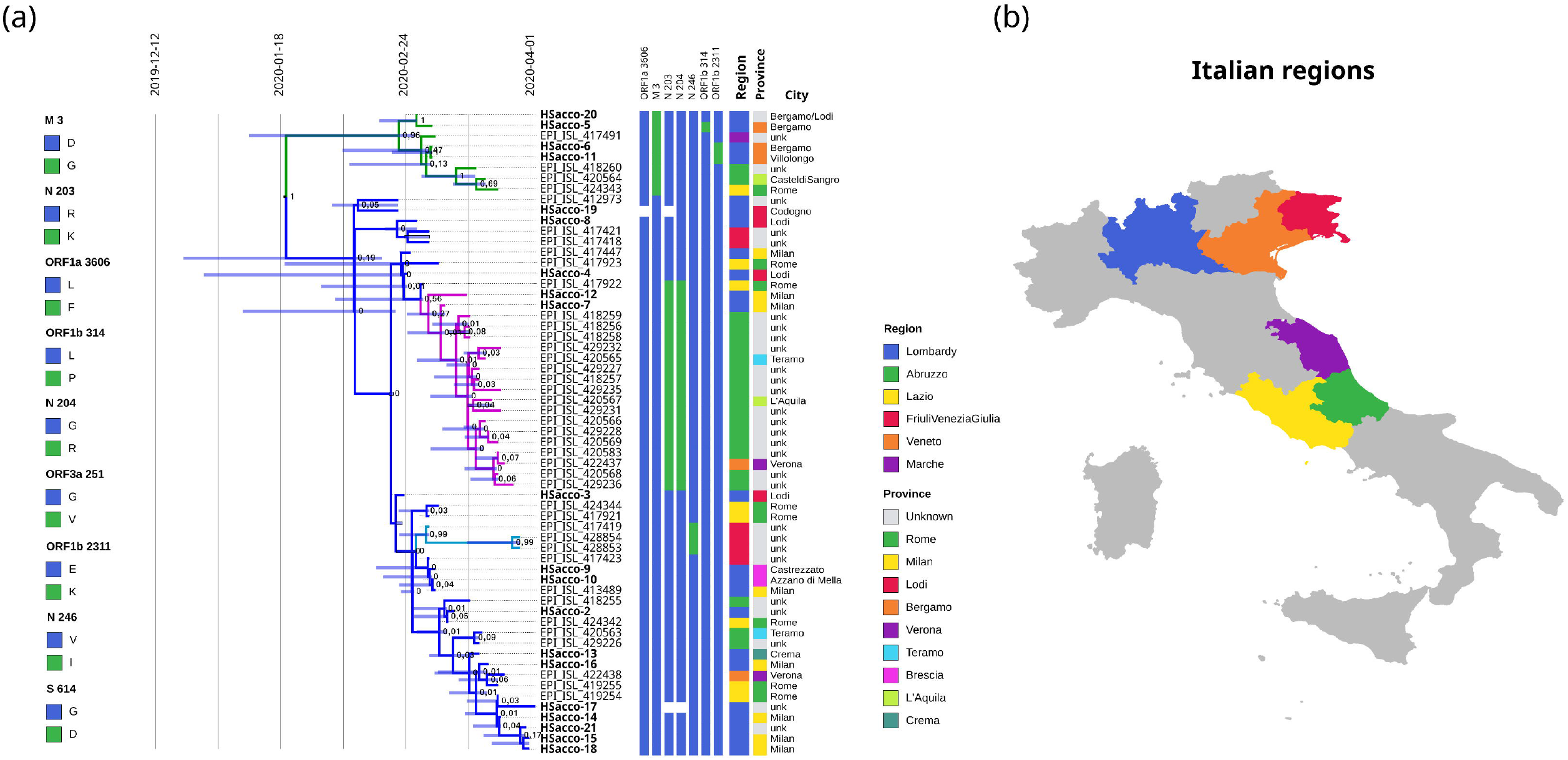

The tree shows the existence of two major monophyletic clades, containing 9 and 52 isolates respectively (coloured in green and blue, respectively, in Figure 1). The smaller clade is characterized by a non-synonymous mutation in position three of the gene M (mutation D3G) and its time of the Most Recent Common Ancestor (tRMCA) resulted 20^th^ February 2020 (*HPD 95%* interval from 4^th^ January to 24^th^ February); the Bergamo and HSacco-20 sequences mapped inside this group. The larger clade contains two monophyletic sub-clades, characterized by non-synonymous mutations in the N gene (Figure 1). The first cluster includes 19 isolates, mainly from Central Italy, in which R203K and G204R variants were found; the other one, instead, accounted for three sequences from Friuli Venezia Giulia region, presenting the V246I substitution. The tMRCA calculated for the two sub-clades were 2^nd^ March 2020 (HPD 95% interval from February 24^th^ to March 6^th^) and 28^th^ February 2020 (HPD 95% interval from February 20^th^ to March 2^nd^), respectively (Table 2).

## DISCUSSION

The Lombardy region has the highest prevalence of COVID-19 in Italy, thus being the likely epicentre of country outbreak. However, it is still unclear how SARS-CoV-2 circulation started: no epidemiological link was found for the first identified autochthonous patient. Moreover, despite the diagnosis was made on 20 February 2020, emerging evidences suggest a multiple virus introduction at least in January 2020. ^21,22,23^

The present work tried to expand understanding on the circulation of SARS-CoV-2 during the early epidemic period in Lombardy, especially in most affected areas. A phylogenetic analysis was conducted on 20 full length genomes collected from patients addressing to several Lombard hospitals from February 20 to April 4, 2020, creating a dataset including 41 Italian viral genome assemblies available on GISAID database as of 30th March, 2020. In spite of restricted geographical origin, these sequences were not included in the same cluster, but two main monophyletic clades containing isolates collected across different regions were found; in addition, the main clade accounting for 52/61 assemblies was divided in two subclades. Interestingly, the molecular clock analysis estimated a clusters divergence approximately one month before the first patient identification. Such evidences are in accordance with other studies, further supporting the hypothesis that different SARS-CoV-2 strains spread all over the world at different time, but their presence became evident only in late February along with Italian epidemic emergence.^21,24,25^ Noteworthy, Bergamo (HSacco-5, HSacco-6, HSacco-11) and HSacco-20 sequences had the M gene D3G mutation of the first clade, in contrast with all the other ones located in the most represented clade. Two main observations consequently arose: firstly, the virus found in Bergamo area seemed to had a more restricted circulation, probably for a delayed introduction. In addition, the nurse working in Lodi and living in Bergamo likely acquired the infection in a personal contest rather than hospital environment, since Lodi patients mapped within the other cluster.

Another interesting outcome was the presence of the N gene mutations R203K and G204R subclade. These two aminoacid changes appear to coexist, since they were always found together into the dataset, as well as they both are present almost exclusively in the GISAID 20B clade. These mutations have an impact on structure and function of the mutated N protein. The phylogenetic tree also clearly showed that the subclade is predominantly made up of isolates from Abruzzo region, suggesting a segregation of this specific virus in Central Italy area. Given that the role of these mutations is still unclear, an Indian group investigated their possible influence on virus replication interference mediated by miRNA: they found out how some miRNA, present in different pathological conditions, are likely to bind to native N gene and repress its expression, thus helping in disease progression limitation; on the contrary, mutated variants could increase their chances of interference escape.^26^

Another small subclade was found in the main one, characterized by the presence of N gene mutation V246I in three sequences, all from Friuli Venezia Giulia region. Besides the unique geographical origin, it is noticeable that in GISAID map ‘Geography’ the V246I mutation is actually present only in Italy, as well as the V246A one only in Israel. Their rarity could have two probable explanations: on the one hand, available data are still limited, making difficult to have a reliable distribution; on the other hand, these variations could have a negative influence on viral fitness, diminishing efficacy in replication and consequently virus transmission.

Other mutations found in the present work had negligible influence on phylogenetic analysis, even a biological significance can not be excluded: viral genome and proteins are key factors in patients management and any variation can extremely burden the efficacy of drugs, vaccines and diagnostic tools or be related to a more severe clinical presentation.^27,28^

In conclusion, this study gave insights on early dynamics of SARS-CoV-2 circulation in Italy, underling peculiar strains localization and supporting multiple virus introductions at least in January 2020.

## Data Availability

SARS-COV-2 sequences are submitted to GISAID; we are waiting for the sequence numbers

